# The relationship between maternal glucose concentrations, gestational diabetes mellitus, placental weight, and placental vascular malperfusion lesions: a retrospective study of a U.S. pregnancy cohort

**DOI:** 10.1101/2025.05.14.25327646

**Authors:** Amrita Arcot, Kelly Gallagher, Jeffery A. Goldstein, Alison D. Gernand

## Abstract

**Background:** Gestational diabetes mellitus (GDM) is associated with increased placental weight and the presence of placental malperfusion lesions, likely related to high blood glucose. The relationship between high glucose without overt GDM, and placental characteristics is not well understood.

**Objective:** To examine the relationships between glucose challenge test (GCT) concentrations, GDM, and placental characteristics associated with GDM.

**Methods:** We conducted a secondary analysis of medical record data from singleton placentas sent to pathology at Northwestern Memorial Hospital (2011-2022; n=11,585). Data included maternal demographic variables, GCT concentrations, GDM diagnosis, placental weight, and vascular malperfusion lesions (accelerated villous maturation, increased syncytial knots, delayed villous maturation, and increased perivillous fibrin deposition). We classified GCT <140 mg/dL as pass and ≥140 mg/dL as fail. We categorized glucose groups into *pass GCT/no GDM*, *fail GCT/no GDM*, and *GDM*. We used linear and Poisson regression models to examine the association between GCT concentrations or groups with placental outcomes, adjusting for maternal age, race and ethnicity, parity, gestational age at delivery, and infant sex.

**Results:** Of placentas sent to pathology, 5% were from pregnancies with GDM and 17% from those who failed the GCT but did not get diagnosed with GDM. Compared to the *pass GCT/no GDM* group, the adjusted mean placental weight was heavier by 13.6 grams [95% CI: 8.8, 18.3] in the *fail GCT/no GDM* and 22.0 grams [13.8, 30.2] in the *GDM* group. Patients diagnosed with GDM had a 36% [2%, 81%] increased adjusted risk of delayed villous maturation compared to the *pass GCT/no GDM*. The risk of the other lesions was not statistically significantly different between groups.

**Conclusion:** GDM and high glucose concentrations without GDM were associated with heavier placentas; patients with GDM had a higher risk of delayed villous maturation.

## Introduction

Gestational diabetes mellitus (GDM) is defined as hyperglycemia or glucose intolerance with first occurrence during pregnancy, and no history of type 1 or type 2 diabetes mellitus [1]. The prevalence of GDM in the United States and Canada is estimated to be 6.9% [2]. In 2021, 16.9 million pregnancies were classified with GDM globally [2]. GDM can result in several adverse pregnancy outcomes, including miscarriage, prematurity, and Cesarean section [3–6]. Of note, GDM typically resolves at delivery, yet five years after pregnancy, the risk of type 2 diabetes mellitus is increased by seven-fold in the mother [7]. Additionally, exposure to hyperglycemia in utero is associated with fetal hyperinsulinemia [8], which could result in insulin resistance in childhood [9]. Consequently, GDM can have acute and long-term health implications for the pregnant person and child.

The placenta supports the growth and development of the fetus [10]. Past studies consistently report increased placental weight in GDM pregnancies, when compared to non-GDM pregnancies [11–45]. Additionally, histological analysis has found an increase in accelerated villous maturation, increased syncytial knots, delayed villous maturation, and increased perivillous fibrin deposition in GDM, type 1 diabetes, and/or type 2 diabetes mellitus pregnancies [25,46–50]. GDM is linked with increased maternal and fetal vascular malperfusion, which includes the above lesions and villous infarcts, fibrinoid necrosis, villous edema, and other complications [51–54]. The mechanisms driving these changes are not well understood and are related, in part, to high maternal blood glucose.

Past literature has established a relationship between GDM and some placental changes, namely weight, but these comparisons are often limited to binary groups of non-GDM controls and overt GDM cases [11–45]. The relationship between placental changes and glucose concentrations on a continuum has not been examined to our knowledge. One method of diagnosing GDM is a two-step strategy: a non-fasted 50-gram glucose challenge test (GCT; screening measure), which, if failed, proceeds to an oral glucose tolerance test (OGTT; diagnostic measure) [55]. Notably, a pregnant person may fail the GDM screening but pass the GDM diagnostic, often referred to as pre-GDM or one abnormal value. Two studies have examined placental outcomes in pregnancies with pre-GDM [56,57]. *Nataly et al.* reported higher proportions of fetal vascular malperfusion lesions in patients with pre-GDM compared to patients with overt GDM [57]. *Rudge et al.* reported a higher proportion of syncytial knots in patients with pre-GDM than in patients with normoglycemia [56]. Additionally, patients with pre-GDM had higher proportions of intervillous fibrosis and delayed villous maturation compared to patients with normoglycemia or overt GDM, albeit not statistically significant. Both studies provide valuable insights into the relationship between glucose and the placenta; however, gaps remain. *Nataly et al.* did not include a general obstetric population as a control.

*Rudge et al.* examined placental samples from patients with normoglycemia (n=6/131), pre-GDM (n=34/311), overt GDM (n=8/131), and overt type II diabetes (n=83/131); notably, the samples comprising normoglycemia and GDM are small. Assessment in a large obstetric sample, with a non-GDM control group, is warranted to examine associations between glucose concentrations, GDM, and placental characteristics.

We aimed to examine the relationships between GCT concentrations, GDM, and placental characteristics, including placental weight and placental vascular malperfusion lesions: accelerated villous maturation, increased syncytial knots, delayed villous maturation, and increased perivillous fibrin deposition. We hypothesized that pregnant patients with high GCT concentrations and those who fail their GCT but “pass” GDM diagnostic test will have a similar placental phenotype to pregnant patients diagnosed with overt GDM.

## Methods

We conducted a retrospective cohort study of anonymized medical record data from Northwestern Memorial Hospital (Northwestern University; Chicago, Illinois) between January 1, 2011 to December 31, 2022. One study investigator (JAG) conducted all data extraction; no other authors had access to or could identify participants during or after data collection.

Northwestern Memorial Hospital is a tertiary-level hospital that performs approximately 10,000 deliveries per year, of which about 20% of delivered placentas are sent to pathology. Medical chart extraction was conducted within an existing study (IRB: STUDY00020697).

### Patient population

All pregnancies within the medical record extraction period were eligible for inclusion if they had a complete pathology report. Placentas sent to pathology are required to meet a certain set of criteria, according to Northwestern Memorial Hospital guidelines. The Northwestern Memorial Hospital decision tree for placental pathology assessment is available in the *Supplementary Materials* (**S1 Fig**). Briefly, placentas can be sent to pathology if they meet criteria for placental (e.g., infarct, cord knot, etc.), fetal (premature < 34 weeks, stillborn, etc.), maternal (severe PE, preterm premature rupture of membranes < 34 weeks, severe preeclampsia, etc.), and/or newborn (Apgar score ≤ 6 at five minutes, ventilatory assistance > 10 minutes, etc.) abnormalities. Of note, GDM is not a criterion for a placental pathology assessment.

Along with a complete pathology report, patients were required to meet the following criteria: singleton pregnancy, completed GDM screening (i.e., GCT available), and gestational age at birth ≥ 140 days (20 weeks, to exclude miscarriage). The exclusion criterion was a history of diagnosed type 1 or type 2 diabetes mellitus.

Our initial sample included 15,675 pregnancies (**Fig 1**). A total of 2,861 were removed per our eligibility criteria, 1,003 were removed due to missingness, and 226 were removed due to biologically implausible values (i.e., placental weight, maternal age, GCT). The final analytic sample was 11,585. We conducted power analysis to determine the power for multivariate analysis given our sample size (n=11,585), for linear, logistic, and Poisson regression [58]. After accounting for at least ten covariates and one degree of freedom, we were powered at 100% for multivariate linear, log-binomial, and Poisson regression, at a significance level of alpha=0.05 and a large effect size. Models and details on the statistical plan are in *Statistical Analysis*.

**Figure 1.**
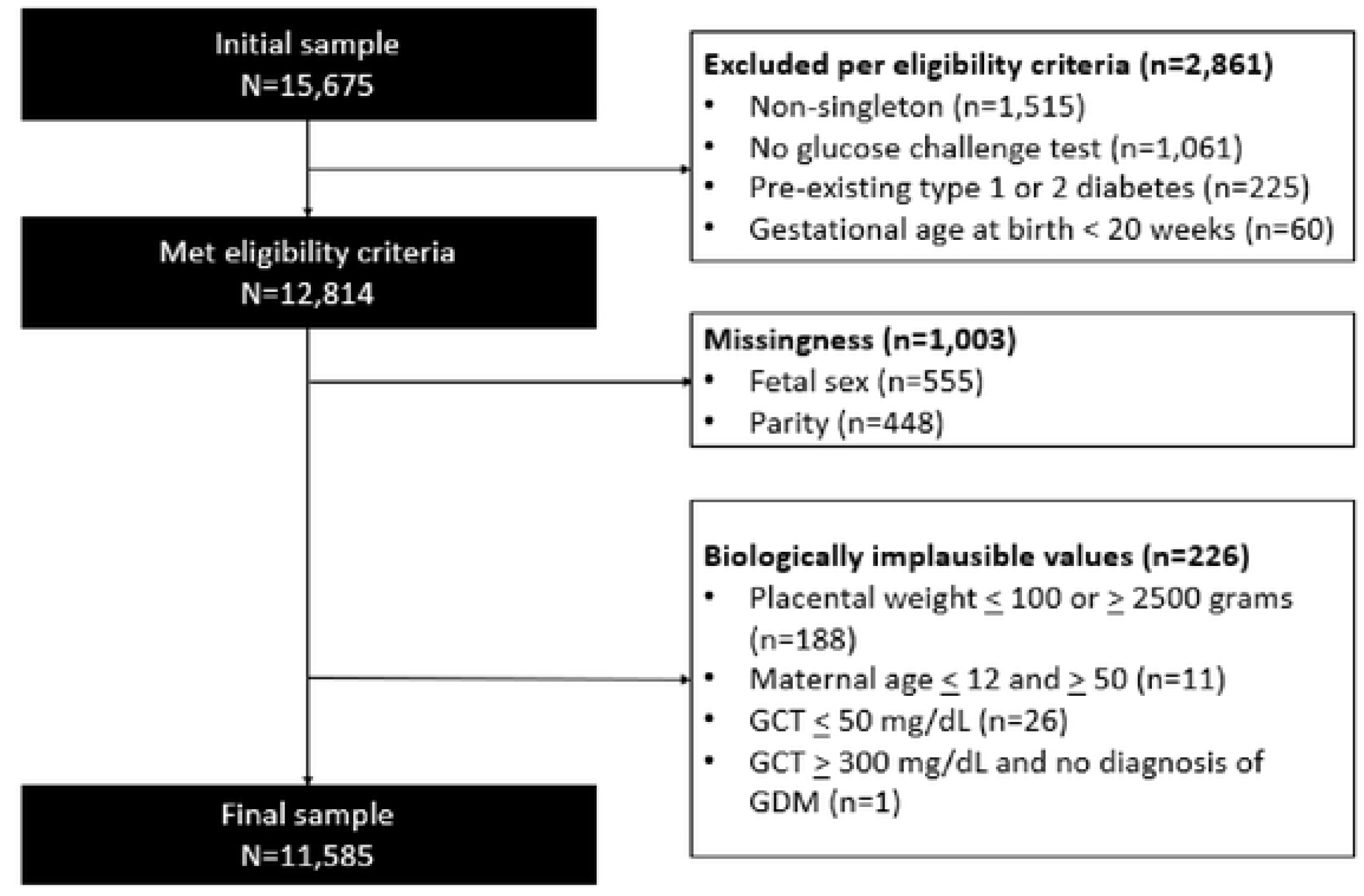
Study selection of pregnant patients from Northwestern Memorial Hospital Abbreviations: GDM=gestational diabetes mellitus

### Covariates

Demographic variables of interest included maternal age, race and ethnicity (see below), parity, gestational age at delivery, infant sex, and maternal hypertension. Maternal age was reported in whole years, parity was patient-reported as the number of births before the current pregnancy, gestational age at delivery was recorded per hospital procedures (last menstrual period and/or first-trimester ultrasound scan), and infant sex was recorded as male, female, or unknown (we recoded as missing as there was no distinction as intersex or other). We classified maternal hypertension based on the International Classification of Diseases, 10^th^ Revision, Clinical Modification (ICD-10-CM) diagnosis codes in the patient’s medical record (**S1 Table**) [59]. The medical record categorized race into two variables: their first identified race (Race 1), and their second identified race (Race 2). These variables were recategorized into the following: Asian (Asian, Asian Indian, Chinese, Filipino, Japanese, Korean, and Other Asian), Black or African American, White, Other (American Indian or Alaska Native, Native Hawaiian, Native Hawaiian or other Pacific Islander, Other Pacific Islander), and Unknown (Blank, Declined, None of the above, Patient declined to respond, Unable to answer, Unknown). The ethnicity variable included the following options: Not Hispanic, Hispanic, Declined, or Patient Unable to Respond. Multiple subcategories were available for those who identify as Hispanic (e.g., “Yes, Puerto Rican”, “Yes, Cuban”, etc.). If a participant reported Declined or Patient Unable to Respond for ethnicity, we recategorized it to ’unknown ethnicity.’ For our analysis, we classified race and ethnicity based on the Race 1 and Ethnicity variables as follows: Non-Hispanic (NH) White (includes White with unknown ethnicity), NH Black or African American (includes Black or African American with unknown ethnicity), NH Asian (includes Asian with unknown ethnicity), Hispanic or Latino (any races), Other, and Unknown. Other includes American Indian and Alaska Native (NH or unknown ethnicity) and Native Hawaiian or Pacific Islander (NH or unknown ethnicity), both of which make up < 1% of the total analytic sample. Unknown was used if a patient did not report their race and/or ethnicity (e.g., declined, unable to answer, etc.). The Race 2 variable was not included because < 1% had a designation (n=73). Infant birth weight was extracted from the medical record as well. We did not have access to maternal height, pre-pregnancy or pregnancy weights, or body mass index (BMI; kg/m^2^) data.

### Glucose and GDM

We identified patients with GDM based ICD-10-CM diagnosis codes in the medical record, detailed in our Supplementary Materials (**S2 Table**) [59]. We classified GDM: (1) GDM, diet control, (2) GDM, insulin control, (3) GDM, oral drug control, and (4) GDM, unspecified control. We grouped all categories to determine the total diagnosis of GDM for the entire study population. Diagnostic terms were not mutually exclusive and total diagnosis of GDM was determined if any of the above categories were present in an individual patient.

Our first exposure of interest was the glucose concentrations from GCT for every patient, with and without GDM, on a continuum. The GCT is a non-fasted screening test, or step one of a two-step strategy, to determine if a patient requires a formal OGTT (diagnostic of GDM) [55]. It consists of a 50-gram glucose drink followed by a blood sample taken one hour post consumption. Our second exposure of interest was three “glucose groups” created based on a patient’s (1) GCT results and (2) diagnosis of GDM (**Table 1**). These groups were created to capture a middle group between non-GDM and GDM that had high glucose in the GCT screening but were not diagnosed with GDM. Of note, 88 participants had a “passing” GCT value but were diagnosed with GDM, likely related to clinical decisions not captured in the medical record data extracted. Those individuals were grouped with participants who were diagnosed with GDM to comprise the *GDM* group.

**Table 1.** Glucose groups based on GCT concentration and GDM diagnosis.

| Groups | GCT result (mg/dL) | Diagnosed with GDM? |
| --- | --- | --- |
| Pass GCT/No GDM | < 140 | No |
| Fail GCT/No GDM | ≥ 140 | No |
| GDM | ≥ 140<sup>†</sup> | Yes |

<sup>†</sup> n=88 had a GCT result < 140 mg/dL but had a GDM diagnosis and were therefore classified in the GDM group. Abbreviations: GCT=glucose challenge test; GDM=gestational diabetes mellitus

Two patients had a questionably high GCT (> 300 mg/dL). The first patient had a high GCT result (398 mg/dL) and did not have a GDM diagnosis, which suggested an error and was deemed biologically implausible. All models were run with and without this one erroneous value with nearly similar results. The investigative team discussed and concluded that this one record (GCT = 398 mg/dL) would be excluded from the primary analysis. The second patient had a high GCT (376 mg/dL) and a GDM diagnosis, which we considered plausible. This patient was kept in the primary analysis, and we conducted a sensitivity analysis excluding the second patient.

### Placental pathology report

We extracted placenta data from the patient’s placental pathology report; perinatal pathologists write all reports following a standardized protocol per Northwestern Memorial Hospital guidelines. Outcomes were placental weight (weighed in the gross exam after trimming back membranes and cutting the umbilical cord < 1 cm from the disc), accelerated villous maturation, increased syncytial knots, delayed villous maturation, and increased perivillous fibrin deposition. Accelerated villous maturation, increased syncytial knots, and delayed villous maturation are defined per the Amsterdam Placental Workshop Group [60]. Perivillous fibrin deposition (also known as massive perivillous fibrin deposition) is characterized by *Faye-Petersen and Ernst* [61]. Medical records may have varied terms for perivillous fibrin deposition. Per the guidance of our placental pathologist (JAG), we combined four lesion diagnoses to characterize increased perivillous fibrin deposition: increased perivillous fibrin, syndromes of perivillous fibrin deposition, massive perivillous fibrin deposition, and borderline massive perivillous fibrin deposition. Our investigative team selected these lesions as they are commonly found in placentas with diabetes [25,46–50].

### Statistical analysis

We used kernel density plots with a normal curve overlay to visualize the normality of distributions for continuous variables. Median, skewness, and kurtosis were examined as well. Placental weight was normally distributed and GCT was close to normal and examined without log transformation [62]. We examined frequencies of discrete variables. We looked for a non-linear relationship between the continuous exposure (GCT) and continuous outcome (placental weight) using LOWESS (Locally Weighted Scatterplot Smoot) plots and found that it was linear.

We assessed demographic characteristics by glucose groups with one-way ANOVA and pairwise t-test for continuous variables with normal distributions (maternal age) and Kruskal-Wallis rank sum test and Dunn’s pairwise test for continuous variables with non-normal distributions (gestational age at birth (days)) [63–65]. We used Pearson’s chi-squared test and proportional pairwise test for categorical variables (race and ethnic group, parity, infant sex, preterm birth) [66,67]. We conducted all pairwise tests with Bonferroni correction [68].

We used linear regression models to estimate the unadjusted and adjusted association of glucose groups and GCT concentrations (each exposure in a separate model) with placental weight. We planned to estimate the unadjusted relative risk (RR) and adjusted relative risk (ARR) for the association of GCT values and glucose groups with the categorical placental outcomes (accelerated villous maturation, increased syncytial knots, delayed villous maturation, and increased perivillous fibrin deposition) with log-binomial regression models; however, models would not converge and Poisson regression with robust standard errors was used instead [69,70]. All models were adjusted for maternal age, maternal race and ethnicity, parity, gestational age at birth, and infant sex. For all adjusted analyses, we collapsed race and ethnicity into three groups (NH White, NH Black or African American, and all other groups) and parity into two groups (0/<u>></u>1).

We examined interactions in these associations by infant sex (male/female) and parity (0/<u>></u>1) and considered an interaction with a *p* value < 0.1 to be statistically significant. For interactions with glucose groups (2-category variable x 3-category variable), we used a Wald test to calculate the *p* value of the combined interaction terms post-estimation [71–73].

In sensitivity analysis, we examined all unadjusted and adjusted models without patients diagnosed with hypertension (defined in **S1 Table**), to test the influence of hypertension on our results.

All statistical tests were two-sided, and we considered results statistically significant if a *p* value was < 0.05. All statistical analyses were run in R, version 4.4.1 [74].

## Results

**Table 2** presents the population characteristics according to different glucose groups. Mean participant age and gestational age at delivery were similar across groups, but statistically different. Participants predominantly self-identified as NH White and were delivering for the first time (parity = 0). Mean glucose concentrations from the GCT were higher than the *pass GCT/no GDM* group, by 60 mg/dL. Just over half of the patients delivered male infants. Infant birthweight was highest in the *pass GCT/no GDM* group, but not statistically different across groups. Mean gestational age was highest in the *pass GCT/no GDM* group. Preterm birth (< 259 days) was proportionally highest in the *fail GCT/no GDM* group.

**Table 2.**
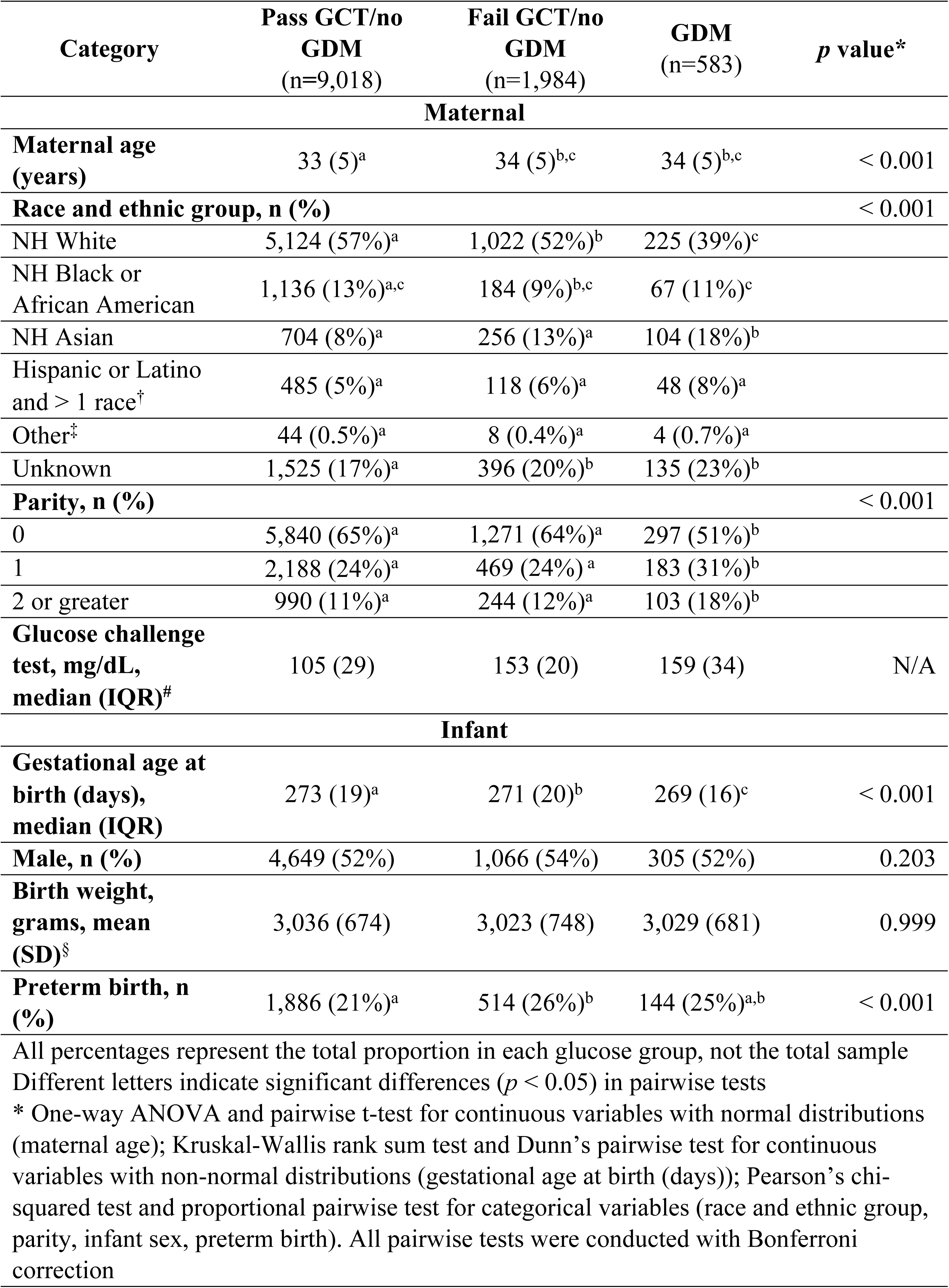
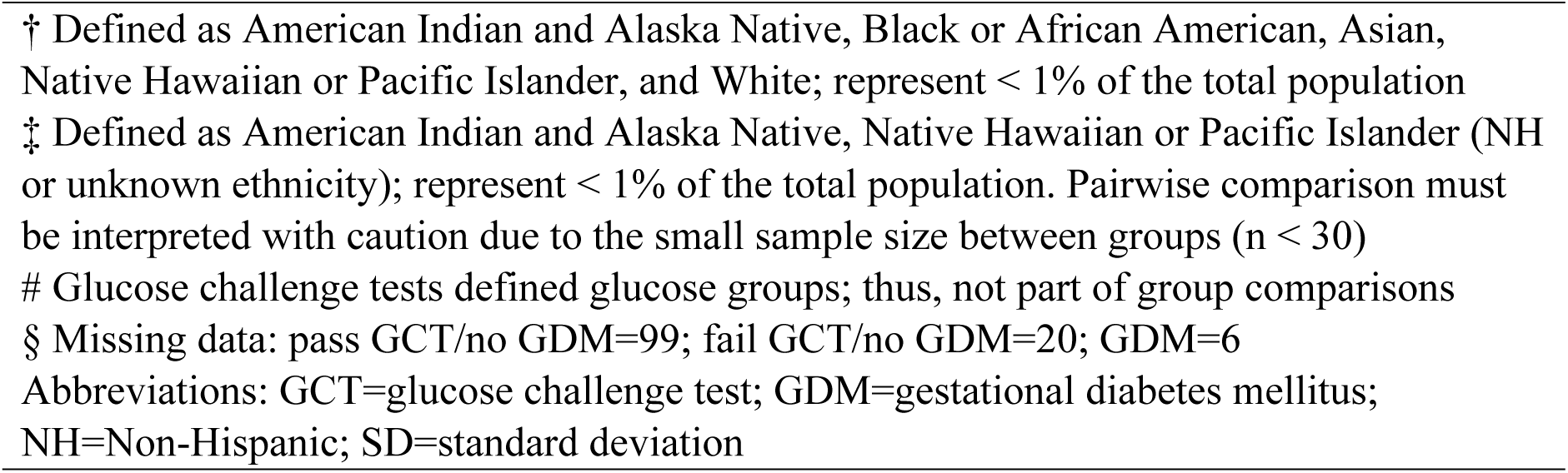
Maternal and infant characteristics by glucose groups (n=11,585)

| Category | Pass GCT/no GDM<br>(n=9,018) | Fail GCT/no GDM<br>(n=1,984) | GDM<br>(n=583) | *p* value* |
| --- | --- | --- | --- | --- |
| **Maternal** |  |  |  |  |
| **Maternal age (years)** | 33 (5)<sup>a</sup> | 34 (5)<sup>b,c</sup> | 34 (5)<sup>b,c</sup> | < 0.001 |
| **Race and ethnic group, n (%)** |  |  |  | < 0.001 |
| NH White | 5,124 (57%)<sup>a</sup> | 1,022 (52%)<sup>b</sup> | 225 (39%)<sup>c</sup> |  |
| NH Black or African American | 1,136 (13%)<sup>a,c</sup> | 184 (9%)<sup>b,c</sup> | 67 (11%)<sup>c</sup> |  |
| NH Asian | 704 (8%)<sup>a</sup> | 256 (13%)<sup>a</sup> | 104 (18%)<sup>b</sup> |  |
| Hispanic or Latino and > 1 race<sup>†</sup> | 485 (5%)<sup>a</sup> | 118 (6%)<sup>a</sup> | 48 (8%)<sup>a</sup> |  |
| Other<sup>‡</sup> | 44 (0.5%)<sup>a</sup> | 8 (0.4%)<sup>a</sup> | 4 (0.7%)<sup>a</sup> |  |
| Unknown | 1,525 (17%)<sup>a</sup> | 396 (20%)<sup>b</sup> | 135 (23%)<sup>b</sup> |  |
| **Parity, n (%)** |  |  |  | < 0.001 |
| 0 | 5,840 (65%)<sup>a</sup> | 1,271 (64%)<sup>a</sup> | 297 (51%)<sup>b</sup> |  |
| 1 | 2,188 (24%)<sup>a</sup> | 469 (24%)<sup>a</sup> | 183 (31%)<sup>b</sup> |  |
| 2 or greater | 990 (11%)<sup>a</sup> | 244 (12%)<sup>a</sup> | 103 (18%)<sup>b</sup> |  |
| **Glucose challenge test, mg/dL, median (IQR)<sup>#</sup>** | 105 (29) | 153 (20) | 159 (34) | N/A |
| **Infant** |  |  |  |  |
| **Gestational age at birth (days), median (IQR)** | 273 (19)<sup>a</sup> | 271 (20)<sup>b</sup> | 269 (16)<sup>c</sup> | < 0.001 |
| **Male, n (%)** | 4,649 (52%) | 1,066 (54%) | 305 (52%) | 0.203 |
| **Birth weight, grams, mean (SD)<sup>§</sup>** | 3,036 (674) | 3,023 (748) | 3,029 (681) | 0.999 |
| **Preterm birth, n (%)** | 1,886 (21%)<sup>a</sup> | 514 (26%)<sup>b</sup> | 144 (25%)<sup>a,b</sup> | < 0.001 |

All percentages represent the total proportion in each glucose group, not the total sample
Different letters indicate significant differences ( $p < 0.05$ ) in pairwise tests
\* One-way ANOVA and pairwise t-test for continuous variables with normal distributions (maternal age); Kruskal-Wallis rank sum test and Dunn's pairwise test for continuous variables with non-normal distributions (gestational age at birth (days)); Pearson's chi-squared test and proportional pairwise test for categorical variables (race and ethnic group, parity, infant sex, preterm birth). All pairwise tests were conducted with Bonferroni correction
† Defined as American Indian and Alaska Native, Black or African American, Asian, Native Hawaiian or Pacific Islander, and White; represent < 1% of the total population
‡ Defined as American Indian and Alaska Native, Native Hawaiian or Pacific Islander (NH or unknown ethnicity); represent < 1% of the total population. Pairwise comparison must be interpreted with caution due to the small sample size between groups (n < 30)

### Glucose concentrations and placental outcomes

The range of placental weights was 100 to 1,265 grams. GCT concentrations had a slight positive relationship with placental weight (**Fig 2**). A 10 mg/dL increase in GCT was associated with a 1.63 gram increase in placental weight (95% CI: 0.95, 2.30, *p* value < 0.001). After adjustment, a 10 mg/dL increase in GCT was more strongly associated with placental weight (3.26 gram; 95% CI: 2.66, 3.86), *p* value < 0.001). The relationship between GCT and placental lesions was non-significant across all models (**S3 Table**).

**Figure 2.**
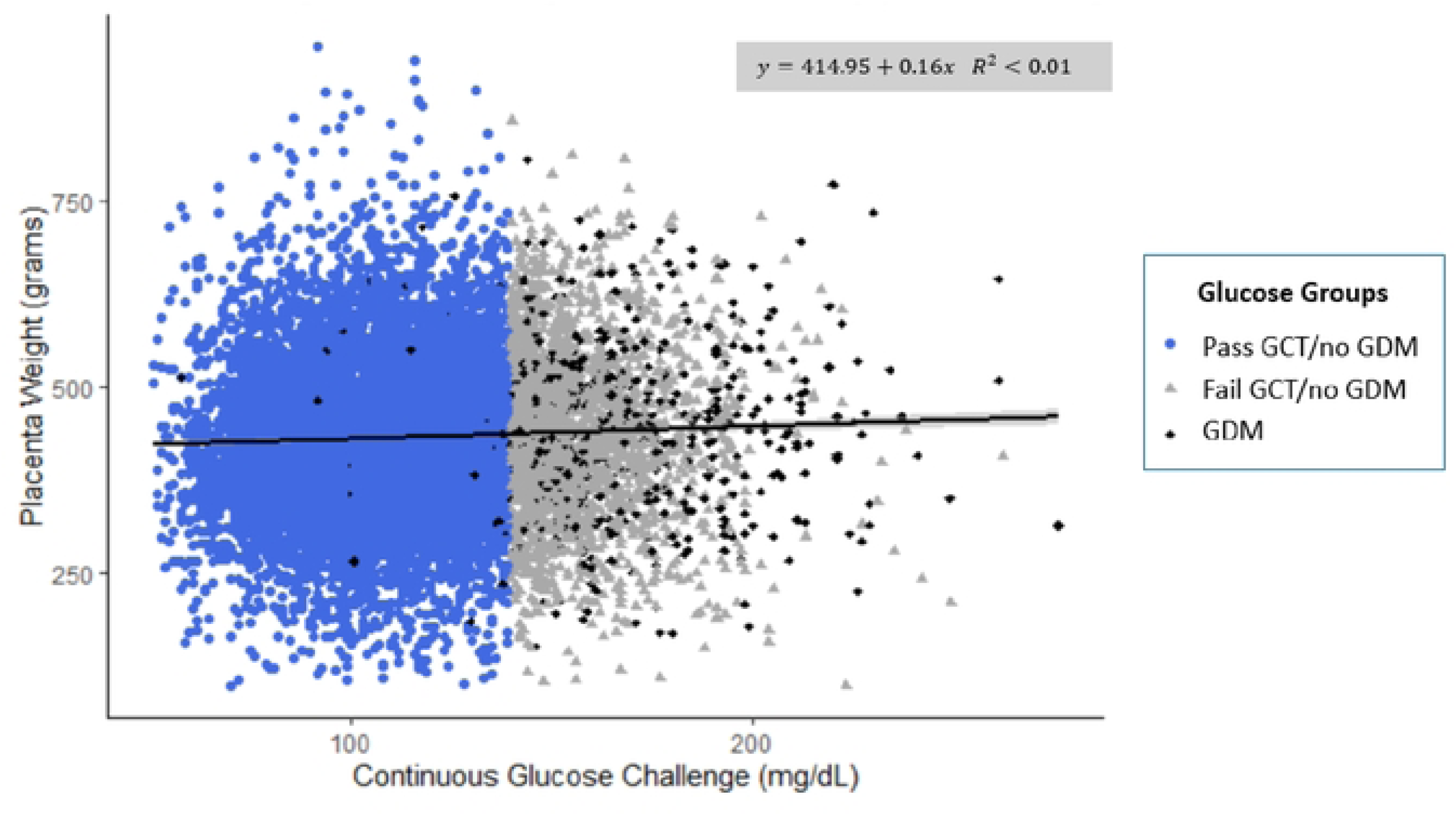
Continuous glucose challenge concentrations (mg/dL) and placental weight (grams) stratified by glucose groups (n=11,583). Excluded from figure: placental weight > 1200 grams (n=1); glucose concentrations > 300 mg/dL (n=1). Abbreviations: GCT=glucose challenge test; GDM=gestational diabetes mellitus

### Glucose groups and placental characteristics

A total of 78% of patients were in the *pass GCT/no GDM* group, 17% were in the *fail GCT/no GDM* group, and the remaining 5% were in the *GDM* group. **S4 Table** shows the frequency of each GDM diagnostic group, with the most prominent diagnosis being *diet control* (n=427, followed by *unspecified control* (n=401). Importantly, diagnoses are not mutually exclusive. Mean (SD) placental weight was highest in the *GDM* group (**Table 3**; 446 grams (111)). Pregnant patients diagnosed with *GDM* had a significantly higher mean difference in placental weight than those in the *pass GCT/no GDM* group (unadjusted mean difference: 14.1 grams (95% CI: 4.9, 23.4; **Table 3**). This difference was higher after adjustment (AMD: 22.0 grams (95% CI: 13.8, 30.2). Pregnant patients in the *fail GCT/no GDM* group had a mean difference in placental weight that was nearly 14 grams higher when compared to the *pass GCT/no GDM* group after adjustment (AMD: 13.6 grams (95% CI: 8.8, 18.3)).

**Table 3.**
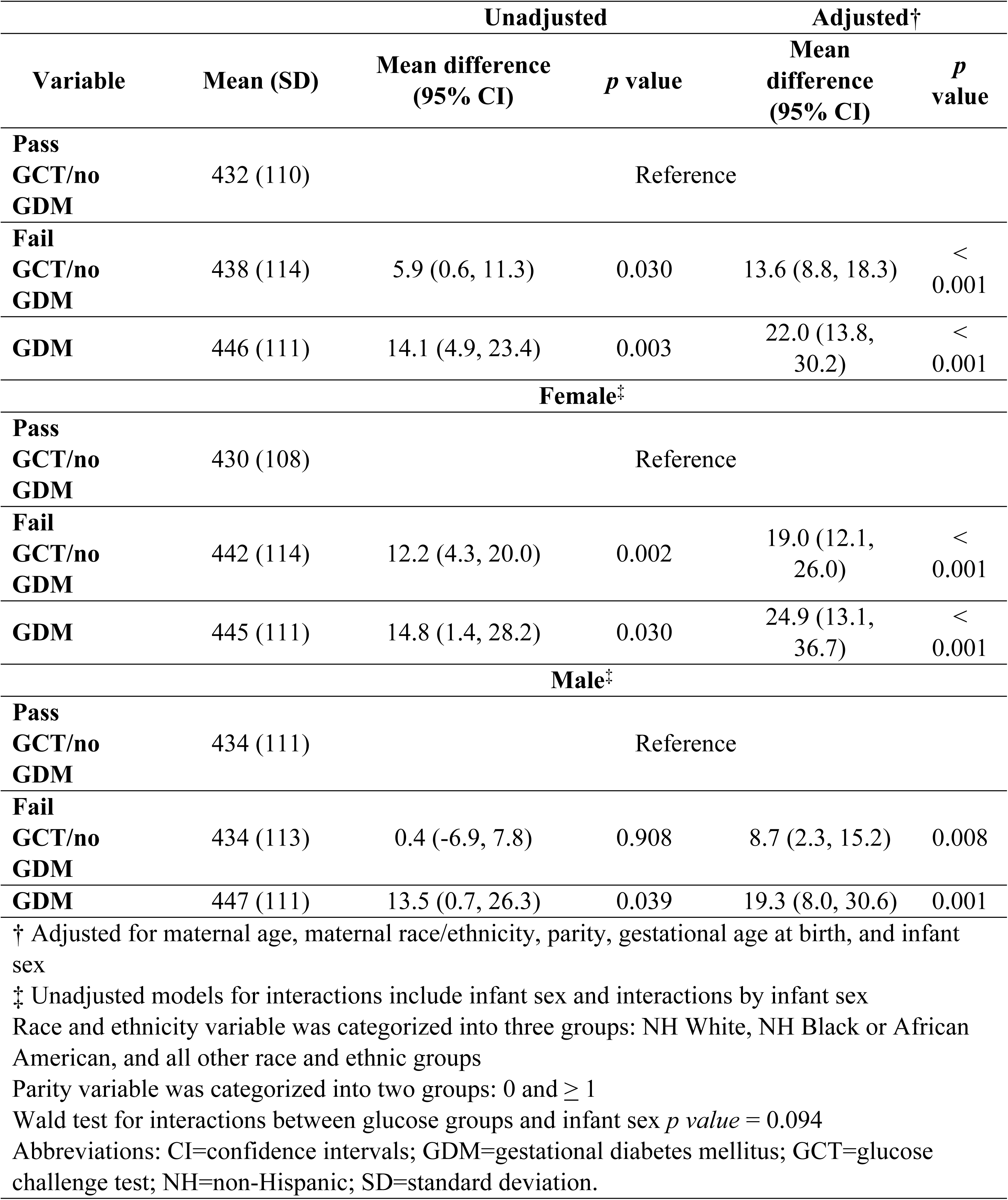
Associations between glucose groups and placental weight (in grams) with interactions by infant sex (n=11,585)

In unadjusted models, accelerated villous maturation risk was 13% (95% CI: 3%, 24%) higher in the fail *GCT/no GDM* group when compared to the *pass GCT/no GDM* group (**S5 Table**). This association was attenuated after adjustment (ARR: 1.00 (0.91, 1.10)). Delayed villous maturation risk was 36% higher in the *GDM* group when compared to the *pass GCT/no GDM* group after adjustment (95% CI: 2%, 81%; **Fig 3**). We found no difference in risk for all other associations of GCT groups and placental lesions.

**Figure 3.**
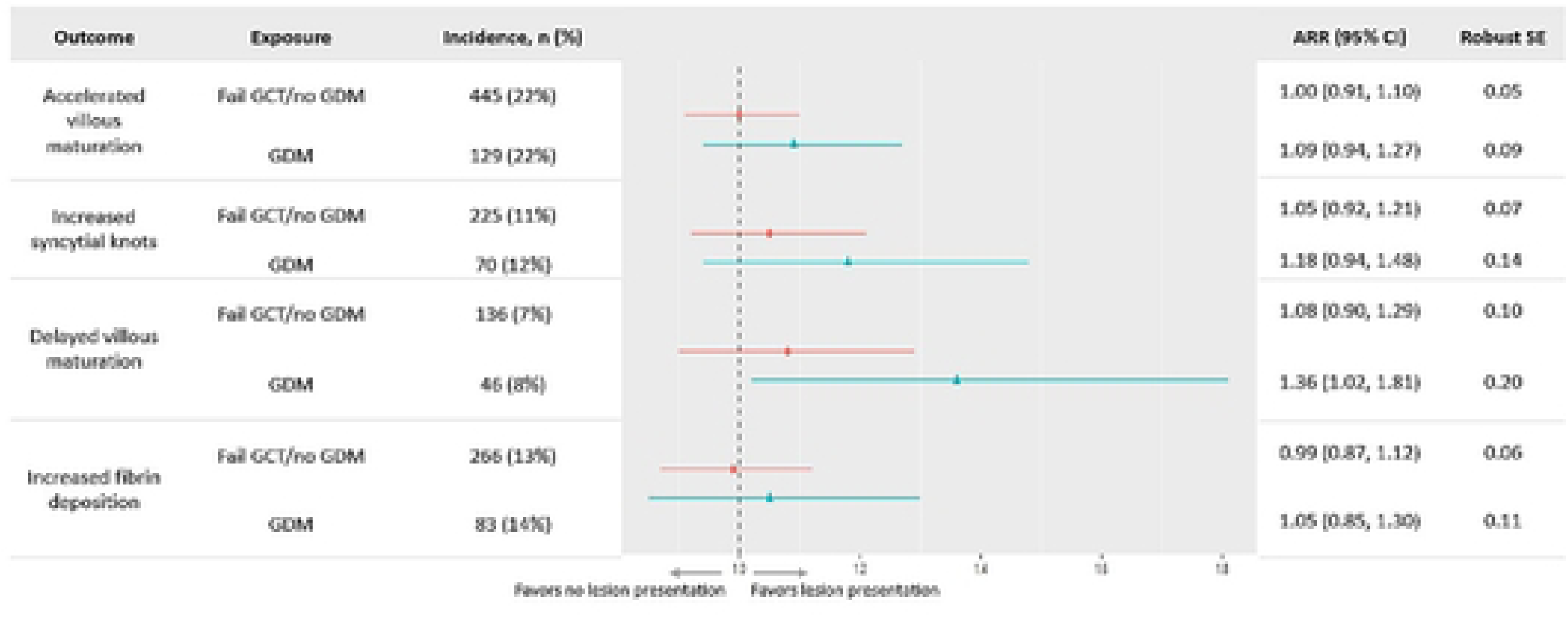
Adjusted relative risk of vascular malperfusion lesions by glucose groups (n=11,585). Circle=Fail GCT/no GDM; Triangles=GDM; Reference glucose group: pass GCT/no GDM; All values were adjusted for maternal age, race, parity, gestational age at birth, and infant sex. Abbreviations: ARR=adjusted relative risk; CI=confidence interval; GCT=glucose challenge test; GDM=gestational diabetes mellitus; NH=non-Hispanic; RR=relative risk; SE=standard error

### Interaction analysis

The relationship between glucose groups and placental weight differed by infant sex (**Table 3**; *p* value = 0.094). Both females and males had heavier placentas in the fail GCT/no GDM and GDM groups, compared to pass GCT/no GDM. For females, the difference between weights in fail GCT/no GDM and pass GCT/no GDM was greater than in males (19.0 vs 8.7). We did not observe an interaction by parity (Wald test *p* value = 0.164).

Sensitivity analysis found no difference in adjusted models for both continuous and categorical exposures when the one elevated GCT (> 300 mg/dL) was excluded. Results were similar for nearly all models when patients with hypertension were excluded (**S6-S9 Tables**). The association between GDM and delayed villous maturation diminished (**S9 Table**), but the spread was similar to the primary analysis.

## Discussion

In this retrospective cohort study of anonymized medical record data from Northwestern Memorial Hospital, we found that GCT on a continuum had a positive relationship with placental weight; however, increased GCT was not associated with an increased risk of placental malperfusion lesions. A failed GCT was associated with higher placental weight in both female and male infants, with and without GDM. Notably, the relationship was strengthened between glucose groups and placental weight after adjustment for covariates. Patients with GDM had a greater risk of delayed villous maturation compared to patients who passed their GCT and were not diagnosed with GDM. Glucose groups were not associated with any other placental malperfusion lesions after adjustment.

Our results reported a very small positive relationship between continuous GCT and placental weight. Both glucose groups were associated with heavier placentas, suggesting that hyperglycemia during pregnancy is positively associated with placental weight, irrespective of a GDM diagnosis. Though small, these differences are within approximately 5% of the average placental weight and are potentially clinically relevant. One past study was identified which examined abnormal glucose values and placental characteristics [57]. *Nataly et al.* conducted a prospective cohort study examining patients with one abnormal value (OAV) in their two-step oral glucose tolerance test (OGTT). Unlike our study, *Nataly et al.* examined OAV in the OGTT (formal diagnostic test for GDM), not the GCT (screen before OGTT). Additionally, they did not include a control group without diagnosed GDM, results were not adjusted for important maternal and infant characteristics associated with placental weight, and they did not conduct interaction analysis by infant sex.

Unlike GCT, the relationship between GDM and increased placental weight has been well-established in past literature [11–45]. *Barke et al.* conducted a case-control study and reported a 44 grams higher unadjusted mean placental weight in GDM pregnancies, though not statistically significant (GDM: 510 (SD: 184) grams versus non-GDM: 466 (SD: 103) grams) [12], much higher than the difference we observed. In a separate case-control study, *Magee et al.* reported a 120 gram higher mean unadjusted placental weight in GDM pregnancies when compared to non-GDM pregnancies (GDM: 570 (SD:10) grams versus non-GDM: 450 (SD: 40) grams) [32]; again, higher than our results. Similar to our research, both studies were conducted in tertiary-level hospitals in the United States and included a non-GDM control group. *Barke et al.* examined placentas sent to pathology per physician discretion, and *Magee et al.* selected participants without pregnancy complications. In contrast to our research, both studies were small (n < 50 for both studies) and did not account for GCT on a continuum [12,32]. Additionally, they did not include an examination of placental malperfusion lesions.

Unlike our findings, some recent literature reported lower mean placental weight in GDM pregnancies than in non-GDM pregnancies [75,76]. Of note, *Hiden et al*. did not test for differences between placental weights (control: 508 (SD=97) grams; GDM: 492 (110) grams) and had a small sample size (n < 30) [75], and *Kadivar et al.* found no statistical difference between mean (SD) placental weights by GDM status (control: 606 (11) grams; GDM: 588 (9) grams) [76], although their difference is similar to the current study. Both studies examined GDM as binary classification, presented unadjusted mean placental weights, and did not examine the relationship between a failed GCT and placental weight. Lastly, *Hiden et al.* examined pregnant patients undergoing cesarean section, and *Kadivar et al.* sampled from a prospective obstetric cohort.

Interaction analysis by infant sex found increased placental weight, if a patient had a failed GCT, in both female and male infants, however, female infants consistently had greater mean placental weight differences compared to males. Unlike females, male placental weights only increased in the *GDM* group, when compared to the *pass GCT/no GDM*. These results suggest that female births may be more sensitive to the maternal environment, compared to male births. Past literature has reported differences in placental size by infant sex, during famine, and periods of fasting (i.e., Ramadan) with contradicting results [77,78]. Studies examining placental efficiency by sex in adverse environments (e.g., asthma, preeclampsia, etc.) have found that females are born smaller than males, but females may have greater placental reserve capacity [79–81]. The mechanisms driving sex-specific differences are not well understood but past evidence suggests greater efficiency in male infants; however, greater placental adaptations are reported in female infants [79,82–84]. Such findings may imply long-term health outcomes in male versus female offspring; however, its relationship with placental weight is unclear.

*Christians and Chow* conducted a large multi-site cohort study (n=2,824) and reported a five-gram greater mean placental weight in males, compared to females, although not significant [85]. Importantly, the investigators did not examine differences by maternal glucose or GDM. Cumulatively, infant sex differences in placental adaptations exist in adverse pregnancy environments but this relationship to placental weight requires further investigation.

The mechanisms driving glucose concentrations and placental weight are not well understood and are largely reported in pregnancies with type 1 diabetes and/or type 2 diabetes [86–88]. *Nelson et al.* reported an increased volume of the intervillous space in pregnancies with type 1 diabetes when compared to controls [86]. *Jauniaux and Burton* reported higher intervillous space volume, as well, along with an increase in trophoblast and placental volume in patients with type 1 diabetes, when compared to healthy controls [87]. Lastly, *Higgins et al.* reported greater terminal villi surface area and immature intermediate villi in pregnancies with type 1 or type 2 diabetes, compared to healthy controls without diabetes [88]. Additionally, capillary surface area was increased in patients with type 1 diabetes compared to the control.

Cumulatively, higher maternal glucose may increase intervillous space volume, placental volume, villi surface area, and capillary surface area. Importantly, all studies were conducted in patients with type 1 and/or type 2 diabetes. These populations should not be generalized to GDM pregnancies as diabetes was present during periconception and the earliest events of placentation. Instead, these findings may provide context for studies examining placental development in GDM pregnancies.

Our results did not find a relationship between high GCT and accelerated villous maturation in adjusted models. Accelerated villous maturation is within the overarching pathology of maternal vascular malperfusion and is characterized by inadequate spiral artery remodeling and thus poor blood flow [54,89]. Past literature does not have a clear consensus on the relationship between accelerated villous maturation and hyperglycemia, partly due to heterogeneity in study designs. *Siassakos et al.* examined placentas sent to pathology and reported an increased incidence of accelerated villous maturation in patients with any abnormal glucose reading in their OGTT (n=6 (50%)) [90]. Interestingly, patients with GDM had a lower incidence of accelerated villous maturation (n=2 (16.7%)). Investigators did not describe the role of dietary and medication intervention in GDM patients. A recent study from *Goto et al.* examining placentas sent to pathology, reported no presence of accelerated villous maturation in GDM placentas, compared to the control [25]. The proportion of maternal vascular malperfusion was greater in the control (n=135 (29%)) when compared to GDM (n=31 (20%), *p* = 0.028). Importantly, *Goto et al.* examined patients who failed their GCT but passed their OGTT. Additionally, neither study examined GCT on a continuum.

Our results also did not find a relationship between high GCT and increased syncytial knots in unadjusted and adjusted models. Syncytial knots are characteristic of accelerated villous maturation and are identified by increased syncytial nuclei at the terminal villi [89,91]. Syncytial knots are an expected pathology with an average proportion of nearly 30% of villi in a term placenta; however, increased syncytial knots can be a sign of immaturity or malperfusion [91]. Increased syncytial knots for gestational age are likely a compensatory mechanism in placental formation to maximize the transfer of nutrients to the fetus [91]. The association between GDM and increased syncytial knots has been well-established in past literature [49,92–95]. *Aldahmash, Alwasel, and Aljerian* reported a greater proportion of increased syncytial knots in pregnancies with GDM (n=34 (77.3%)) compared to the control (n=11 (27.5%); *p* < 0.01) [48]. Similarly, *Dasgupta et al.* reported a greater incidence of increased syncytial knots that were higher in pregnancies with GDM (n=36 (86%)) compared to the control (n=8 (19%); *p <* 0.001) [47]. Both studies were case-control, had similar sample sizes, and defined increased syncytial knots as more than 30-33% of villi in the placenta [60,96]. Of note, *Dasgupta et al.* examined placentas that were sent to pathology – as such pregnancies with GDM were not compared to a healthy, normal obstetric population. In contrast, *Bhattacharjee et al.* conducted a cross-sectional analysis of placentas sent to pathology and reported no significant difference in increased syncytial knots between pregnancies with GDM (n=7 (21.2%)) compared to the control (n=2 (16.7%), *p* = 1.00) [97]. Patients with mild hyperglycemia (defined as normal OGTT with an altered glucose profile) had greater syncytial knots (n=5 (28.5%), *p* = 0.4), albeit non-significant, when compared to the control.

Our results reported an increased risk of delayed villous maturation in pregnancies with GDM when compared to the control. Delayed villous maturation is a supportive finding of fetal vascular malperfusion and is characterized by a reduction in the critical vascular branching of the chorionic villi and thus a reduction in vasculosyncytial membrane formation [52,53,98,99]. A failed GCT without overt GDM was not associated with an increased risk of delayed villous maturation in unadjusted or adjusted models. Past literature supports the relationship between delayed villous maturation and GDM [93,94,97,100,101]. Recent evidence suggests an association between abnormal glucose values, without GDM, and delayed villous maturation. *Nataly et al.* reported an increased incidence of fetal vascular malperfusion lesions in their OAV group (n=25 (20%)), compared to patients with GDM receiving medication therapy (n=15 (25.4%), *p* = 0.02). In contrast, our study found no relationship between GCT on a continuum or a failed GCT without GDM and select fetal malperfusion lesions (delayed villous maturation and increased perivillous fibrin deposition). Of note, *Nataly et al.* examined patients with an abnormal value in their OGTT, whereas our study examined a failed GCT (glucose screen before the OGTT). Findings may suggest further investigation into abnormal values in the OGTT itself and the risk of vascular malperfusion lesions. *Rudge et al.* examined patients with mild gestational hyperglycemia (MGH; defined as a normal OGTT with at least two borderline values) [56]. Placental dysmaturity (not defined) was present in patients with MGH (n=5 (14.7%)), GDM (n=3 (37.5%)), and overt diabetes (n=8 (9.6%)), but not controls. Although non-significant, MGH can lead to the incidence of placental lesions. Importantly, sample comparisons were unbalanced, with six patients with normoglycemia, eight patients with GDM, 34 with MGH, and 83 with ‘overt’ diabetes (not defined). Additionally, the categorization of MGH was not well defined and suggested that patients were required to pass their OGTT and have at least two borderline values. Those who were diagnosed with pre-gestational diabetes were categorized as overt diabetes and not MGH.

Finally, our results found no associations between glucose groups and perivillous fibrin deposition. Perivillous fibrin deposition is characterized by excessive fibrin and trophoblasts surrounding the terminal villi [102]. An increase in fibrin deposition is a criterion for global partial fetal vascular malperfusion, which is linked to obstructions to the umbilical cord [52]. The relationship between glucose concentrations, specifically GDM, and increased perivillous fibrin deposition is inconsistent. This may be partly due to the different locations of fibrin deposition in the placenta (intramural versus intervillous versus perivillous) [25,47,48]. Importantly, these lesions should not be grouped and generalized when examining GDM pregnancies. Dasgupta et al. reported a significantly higher proportion of increased perivillous fibrin deposition in pregnancies with GDM (n=20, 48%) compared to the control (n=10, 24%, *p* =0.023) [47].

Similar to our methods, *Dasgupta et al.* conducted their study in a tertiary-level hospital and examined placentas sent to pathology. Unlike our study, the association between fibrin deposition and failed GCT was not examined.

Strengths of our study include that we extracted and analyzed a large medical record dataset from a high-resource, tertiary-level hospital. We examined placental outcomes from complete pathology reports with expert assistance in interpretation from an experienced placental pathologist (JAG). Additionally, we selected and examined lesions on the maternal *and* fetal sides of the placenta. Our study is not without limitations. All data were extracted from placentas sent to pathology, and thus, comparisons were not conducted with a general obstetric population. The medical records included neither pre-pregnancy BMI nor the ability to calculate it for inclusion in our adjusted analysis. While our large sample size made the study overpowered to detect statistical differences, the statistically significant results were also, in our opinion, meaningfully different and findings on placental weight were consistent across analyses. Despite these limitations, our findings provide foundational knowledge on the relationship between glucose concentrations, placental morphology, and placental lesions.

## Conclusions

Our findings suggest that pregnant patients with elevated glucose concentrations have heavier placentas than those without elevated glucose during pregnancy. Interaction analysis found heavier placentas in females, compared to males, in patients with elevated glucose compared to patients without elevated glucose. These associations strengthened after adjustment. Patients with diagnosed GDM have an increased risk of delayed villous maturation, compared to the pass GCT/no GDM control. The relationships between glucose groups and placental malperfusion lesions (maternal and fetal) were otherwise null. Future studies are necessary to examine other placental morphological characteristics, such as placental diameter, volume, and central thickness. Future studies should also examine the clinical significance of patients who failed their GCT without overt GDM and their chronic disease risk postpartum (e.g., type 2 diabetes mellitus). The placenta is a complex organ that provides a window into pregnancy health, and the present study found that placentas are responsive to high glucose even in patients without a GDM diagnosis. Future research could investigate the potential for placental characteristics (such as weight) to aid in predicting and understanding health outcomes in both the pregnant person and the infant, particularly in settings where glucose testing is not possible or missed.

## Data Availability

Data are available after the execution of a data use agreement with Northwestern University. Investigators should contact Dr. Goldstein.

## Ethics Approval Statement

Medical chart extraction was conducted as part of an existing multi-institution study with ethical approval through a single-IRB protocol at The Pennsylvania State University (IRB: STUDY00020697). Northwestern University was a participating site with a reliance agreement under the single-IRB.

## Author Contributions

All authors contributed to the interpretation and revision of the manuscript. Conceptualization: AA, KG, ADG; Statistical analysis: AA; Writing (original preparation): AA; Writing (reviewing and editing): KG, JAG, ADG. The authors have read and agreed to the published version of this manuscript.

## Supporting information

**S1 Figure. Placental pathology decision tree from Northwestern Memorial Hospital.** Abbreviations: APGAR= appearance, pulse, grimace, activity, and respiration; CMV=cytomegalovirus; DR=delivery room; HSV=herpes simplex virus; IUFD=intrauterine fetal demise; N=no; NICU=neonatal intensive care unit; PPROM=preterm premature rupture of membranes; SGA=small for gestational age; Y=yes

**S1 Table. Maternal hypertension categories by ICD-10-CM.**

All diagnoses were collapsed into the overall category “Maternal Hypertension” Abbreviations: ICD-10-CM= International Classification of Diseases, 10^th^ Revision, Clinical Modification; PE=preeclampsia; HTN=hypertension; HELLP: hemolysis, elevated liver enzymes, and low platelets; w/o=without

**S2 Table. Gestational diabetes mellitus diagnosis by ICD-10-CM.**

All diagnoses were collapsed into the overall category “GDM”

Abbreviations: ICD-10-CM= International Classification of Diseases, 10^th^ Revision, Clinical Modification; GDM=Gestational diabetes mellitus; Gestatnl diab in chldbrth ctrl by oral hypoglycemic drugs=Gestational diabetes in childbirth controlled by oral hypoglycemic drugs

**S3 Table. Associations between glucose challenge tests (per 10 mg/dL increase) and placental lesions (n=11,585).**

† Poisson regression model adjusted for maternal age, race and ethnicity, parity, gestational age at delivery, and infant sex

Abbreviations: ARR=adjusted relative risk; CI=confidence interval; RR=relative risk; SE=standard error

**S4 Table. GDM diagnostic criteria and total frequency.**

† Diagnoses are not mutually exclusive (i.e. some women have >1 GDM diagnosis)

‡ Only from the current pregnancy, history of GDM diagnoses not included Abbreviations: GDM=gestational diabetes mellitus

**S5 Table. Associations between glucose groups and placental lesions (n=11,585).**

The units for glucose challenge tests were 10 mg/dL

Interactions by infant sex and parity were not significant for any models (Wald test ≥ 0.1) and thus not included in this table

† Poisson regression model adjusted for maternal age, race (reference = NH White), parity (reference = 0), gestational age at delivery, and infant sex (reference = Female) Abbreviations: ARR=adjusted relative risk; CI=confidence interval; GCT=glucose challenge test; GDM=gestational diabetes mellitus; RR=relative risk; SE=standard error

**S6 Table. Associations between glucose challenge tests (per 10 mg/dL increase) and placental weight, a sensitivity analysis excluding patients diagnosed with maternal hypertension (n=10,832).**

A total of 753 patients were diagnosed with maternal hypertension and were excluded

† Linear regression models were adjusted for maternal age, race and ethnicity, parity, gestational age at delivery, and fetal sex

Abbreviations: AMD=adjusted mean difference; CI=confidence interval; MD=mean difference

**S7 Table. Associations between glucose challenge tests (per 10 mg/dL increase) and categorical placental lesions (n=10,832).**

A total of 753 patients were diagnosed with maternal hypertension and were excluded

† Poisson regression models were adjusted for maternal age, race and ethnicity, parity, gestational age at delivery, and fetal sex

Abbreviations: ARR=adjusted relative risk; CI=confidence interval; LGA=large for gestational age; RR=relative risk; SGA=small for gestational age

**S8 Table. Associations between glucose groups and placental weight, a sensitivity analysis excluding patients diagnosed with maternal hypertension (n=10,832).**

A total of 753 patients were diagnosed with maternal hypertension and were excluded

† Linear regression model was adjusted for maternal age, race and ethnicity, parity, gestational age at delivery, and fetal sex

Abbreviations: CI=confidence intervals; GDM=gestational diabetes mellitus; GCT=glucose challenge test; NH=non-Hispanic; SD=standard deviation

**S9 Table. Associations between glucose groups and categorical outcomes, a sensitivity analysis excluding patients diagnosed with maternal hypertension (n=10,832).**

† Poisson regression model was adjusted for maternal age, race and ethnicity, parity, gestational age at delivery, and fetal sex

Abbreviations: CI=confidence intervals; GDM=gestational diabetes mellitus; GCT=glucose challenge test

## References

[1] Centers for Disease Control and Prevention, U.S. Department of Health and Human Services. National Diabetes Statistics Report, 2020: Estimates of diabetes and its burden in the United States 2020:32.

[2] Eades CE, Burrows KA, Andreeva R, Stansfield DR, Evans JM. Prevalence of gestational diabetes in the United States and Canada: a systematic review and meta-analysis. BMC Pregnancy Childbirth 2024;24:204. 10.1186/s12884-024-06378-2.

[3] Yang H, Wei Y, Gao X, Xu X, Fan L, He J, et al. Risk factors for gestational diabetes mellitus in Chinese women—a prospective study of 16 286 pregnant women in China. Diabetic Medicine 2009;26:1099–104.

[4] Fadl HE, Östlund IKM, Magnuson AFK, Hanson USB. Maternal and neonatal outcomes and time trends of gestational diabetes mellitus in Sweden from 1991 to 2003. Diabetic Medicine 2010;27:436–41. 10.1111/j.1464-5491.2010.02978.x.

[5] Gorgal R, Gonçalves E, Barros M, Namora G, Magalhães Â, Rodrigues T, et al. Gestational diabetes mellitus: A risk factor for non-elective cesarean section. Journal of Obstetrics and Gynaecology Research 2012;38:154–9. 10.1111/j.1447-0756.2011.01659.x.

[6] Bas-lando M, Srebnik N, Farkash R, Ioscovich A, Samueloff A, Grisaru-Granovsky S. Elective induction of labor in women with gestational diabetes mellitus: an intervention that modifies the risk of cesarean section. Arch Gynecol Obstet 2014;290:905–12. 10.1007/s00404-014-3313-6.

[7] Bellamy L, Casas J-P, Hingorani AD, Williams D. Type 2 diabetes mellitus after gestational diabetes: a systematic review and meta-analysis. Lancet 2009;373:1773–9. 10.1016/S0140-6736(09)60731-5.

[8] The HAPO Study Cooperative Research Group. Hyperglycemia and Adverse Pregnancy Outcomes. N Engl J Med 2008;358:1991–2002. 10.1056/NEJMoa0707943.

[9] Scholtens DM, Kuang A, Lowe LP, Hamilton J, Lawrence JM, Lebenthal Y, et al. Hyperglycemia and Adverse Pregnancy Outcome Follow-up Study (HAPO FUS): Maternal Glycemia and Childhood Glucose Metabolism. Diabetes Care 2019;42:381–92. 10.2337/dc18-2021.

[10] Turco MY, Moffett A. Development of the human placenta. Development 2019;146:dev163428.

[11] Algaba-Chueca F, Maymó-Masip E, Ejarque M, Ballesteros M, Llauradó G, López C, et al. Gestational diabetes impacts fetal precursor cell responses with potential consequences for offspring. Stem Cells Translational Medicine 2020;9:351–63. 10.1002/sctm.19-0242.

[12] Barke TL, Goldstein JA, Sundermann AC, Reddy AP, Linder JE, Correa H, et al. Gestational diabetes mellitus is associated with increased CD163 expression and iron storage in the placenta. American J Rep Immunol 2018;80:e13020. 10.1111/aji.13020.

[13] Berglund SK, García-Valdés L, Torres-Espinola FJ, Segura MT, Martínez-Zaldívar C, Aguilar MJ, et al. Maternal, fetal and perinatal alterations associated with obesity, overweight and gestational diabetes: an observational cohort study (PREOBE). BMC Public Health 2016;16:207. 10.1186/s12889-016-2809-3.

[14] Bianchi C, Taricco E, Cardellicchio M, Mandò C, Massari M, Savasi V, et al. The role of obesity and gestational diabetes on placental size and fetal oxygenation. Placenta 2021;103:59–63. 10.1016/j.placenta.2020.10.013.

[15] Choo S, De Vrijer B, Regnault TRH, Brown HK, Stitt L, Richardson BS. The impact of maternal diabetes on birth to placental weight ratio and umbilical cord oxygen values with implications for fetal-placental development. Placenta 2023;136:18–24. 10.1016/j.placenta.2023.02.008.

[16] Cvitic S, Novakovic B, Gordon L, Ulz CM, Mühlberger M, Diaz-Perez FI, et al. Human fetoplacental arterial and venous endothelial cells are differentially programmed by gestational diabetes mellitus, resulting in cell-specific barrier function changes. Diabetologia 2018;61:2398–411. 10.1007/s00125-018-4699-7.

[17] Davenport MH, Campbell MK, Mottola MF. Increased incidence of glucose disorders during pregnancy is not explained by pre-pregnancy obesity in London, Canada. BMC Pregnancy Childbirth 2010;10:85. 10.1186/1471-2393-10-85.

[18] Díaz-Pérez FI, Hiden U, Gauster M, Lang I, Konya V, Heinemann A, et al. Post-transcriptional down regulation of ICAM-1 in feto-placental endothelium in GDM. Cell Adhesion & Migration 2016;10:18–27. 10.1080/19336918.2015.1127467.

[19] Diceglie C, Anelli GM, Martelli C, Serati A, Lo Dico A, Lisso F, et al. Placental Antioxidant Defenses and Autophagy-Related Genes in Maternal Obesity and Gestational Diabetes Mellitus. Nutrients 2021;13:1303. 10.3390/nu13041303.

[20] Du R, Wu N, Bai Y, Tang L, Li L. circMAP3K4 regulates insulin resistance in trophoblast cells during gestational diabetes mellitus by modulating the miR-6795-5p/PTPN1 axis. J Transl Med 2022;20:180. 10.1186/s12967-022-03386-8.

[21] El Hajj N, Pliushch G, Schneider E, Dittrich M, Müller T, Korenkov M, et al. Metabolic programming of MEST DNA methylation by intrauterine exposure to gestational diabetes mellitus. Diabetes 2013;62:1320–8. 10.2337/db12-0289.

[22] Elamin A, Mohammed Ahmed M, El Elhaj A, Ahmed Hussien T, Abdelrahman Mohamed A, Mohamed H, et al. Vicissitudes in the placental cotyledon number in a singleton pregnancy with gestational diabetes. Int J App Basic Med Res 2022;12:24. 10.4103/ijabmr.ijabmr_230_21.

[23] Ćetković A, Miljic D, Ljubić A, Patterson M, Ghatei M, Stamenković J, et al. Plasma Kisspeptin Levels in Pregnancies with Diabetes and Hypertensive Disease as a Potential Marker of Placental Dysfunction and Adverse Perinatal Outcome. Endocrine Research 2012;37:78–88. 10.3109/07435800.2011.639319.

[24] Gauster M, Desoye G, Tötsch M, Hiden U. The Placenta and Gestational Diabetes Mellitus. Curr Diab Rep 2012;12:16–23. 10.1007/s11892-011-0244-5.

[25] Goto T, Sato Y, Kodama Y, Tomimori K, Sameshima H, Aman M, et al. Association between fetal vascular malperfusion and gestational diabetes. The Journal of Obstetrics and Gynaecology Research 2022;48:80–6.

[26] Jadhav A, Khaire A, Gundu S, Wadhwani N, Chandhiok N, Gupte S, et al. Placental neurotrophin levels in gestational diabetes mellitus. Intl J of Devlp Neuroscience 2021;81:352–63. 10.1002/jdn.10107.

[27] Jeelani H, Jabeen F, Qureshi A, Mushtaq S. Gross morphological alterations and birth weight in gestational diabetes mellitus: a case-control study. JK Practitioner 2015;20:17–20.

[28] Klid S, Algaba-Chueca F, Maymó-Masip E, Guarque A, Ballesteros M, Diaz-Perdigones C, et al. The angiogenic properties of human amniotic membrane stem cells are enhanced in gestational diabetes and associate with fetal adiposity. Stem Cell Res Ther 2021;12:608. 10.1186/s13287-021-02678-y.

[29] Kucuk M, Doymaz F. Placental weight and placental weight-to-birth weight ratio are increased in diet- and exercise-treated gestational diabetes mellitus subjects but not in subjects with one abnormal value on 100-g oral glucose tolerance test. Journal of Diabetes and Its Complications 2009;23:25–31. 10.1016/j.jdiacomp.2007.04.002.

[30] Lao T, Lee C, Wong W. Placental weight to birthweight ratio is increased in mild gestational glucose intolerance. Placenta 1997;18:227–30. doi: 10.1016/s0143-4004(97)90097-7.

[31] Madazli R, Tuten A, Calay Z, Uzun H, Uludag S, Ocak V. The incidence of placental abnormalities, maternal and cord plasma malondialdehyde and vascular endothelial growth factor levels in women with gestational diabetes mellitus and nondiabetic controls. Gynecologic and Obstetric Investigation 2008;65:227–32. 10.1159/000113045.

[32] Magee TR, Ross MG, Wedekind L, Desai M, Kjos S, Belkacemi L. Gestational diabetes mellitus alters apoptotic and inflammatory gene expression of trophobasts from human term placenta. Journal of Diabetes and Its Complications 2014;28:448–59. 10.1016/j.jdiacomp.2014.03.010.

[33] Martino J, Sebert S, Segura MT, García-Valdés L, Florido J, Padilla MC, et al. Maternal Body Weight and Gestational Diabetes Differentially Influence Placental and Pregnancy Outcomes. The Journal of Clinical Endocrinology & Metabolism 2016;101:59–68. 10.1210/jc.2015-2590.

[34] McManus R, Summers K, De Vrijer B, Cohen N, Thompson A, Giroux I. Maternal, umbilical arterial and umbilical venous 25-hydroxyvitamin D and adipocytokine concentrations in pregnancies with and without gestational diabetes. Clinical Endocrinology 2014;80:635–41. 10.1111/cen.12325.

[35] Meng Q, Shao L, Luo X, Mu Y, Xu W, Gao L, et al. Expressions of VEGF-A and VEGFR-2 in placentae from GDM pregnancies. Reprod Biol Endocrinol 2016;14:61. 10.1186/s12958-016-0191-8.

[36] Schliefsteiner C, Peinhaupt M, Kopp S, Lögl J, Lang-Olip I, Hiden U, et al. Human Placental Hofbauer Cells Maintain an Anti-inflammatory M2 Phenotype despite the Presence of Gestational Diabetes Mellitus. Front Immunol 2017;8:888. 10.3389/fimmu.2017.00888.

[37] Segura MT, Demmelmair H, Krauss-Etschmann S, Nathan P, Dehmel S, Padilla MC, et al. Maternal BMI and gestational diabetes alter placental lipid transporters and fatty acid composition. Placenta 2017;57:144–51. 10.1016/j.placenta.2017.07.001.

[38] Taricco E, Radaelli T, Nobile de Santis MS, Cetin I. Foetal and Placental Weights in Relation to Maternal Characteristics in Gestational Diabetes. Placenta 2003;24:343–7. 10.1053/plac.2002.0913.

[39] Taricco E, Radaelli T, Rossi G, Nobile de Santis M, Bulfamante G, Avagliano L, et al. Effects of gestational diabetes on fetal oxygen and glucose levels in vivo. BJOG: An International Journal of Obstetrics & Gynaecology 2009;116:1729–35. 10.1111/j.1471-0528.2009.02341.x.

[40] Tumminia A, Scalisi NM, Milluzzo A, Ettore G, Vigneri R, Sciacca L. Maternal Diabetes Impairs Insulin and IGF-1 Receptor Expression and Signaling in Human Placenta. Front Endocrinol 2021;12:621680. 10.3389/fendo.2021.621680.

[41] Visiedo F, Bugatto F, Sánchez V, Cózar-Castellano I, Bartha JL, Perdomo G. High glucose levels reduce fatty acid oxidation and increase triglyceride accumulation in human placenta. American Journal of Physiology-Endocrinology and Metabolism 2013;305:E205–12. 10.1152/ajpendo.00032.2013.

[42] Strøm-Roum EM, Jukic AM, Eskild A. Offspring birthweight and placental weight—does the type of maternal diabetes matter? A population-based study of 319 076 pregnancies. Acta Obstet Gynecol Scand 2021;100:1885–92. 10.1111/aogs.14217.

[43] Wang C-Y, Su M-T, Cheng H, Kuo P-L, Tsai P-Y. Fetuin-A Inhibits Placental Cell Growth and Ciliogenesis in Gestational Diabetes Mellitus. IJMS 2019;20:5207. 10.3390/ijms20205207.

[44] Marin AJ, Ionescu-Tirgoviste C. Fetal proinsulin and insulin and placental weight in pregnancies complicated by gestational diabetes and obesity. GinecologiaRo 2012;8:140–5.

[45] Pagán A, Prieto-Sánchez MT, Blanco-Carnero JE, Gil-Sánchez A, Parrilla JJ, Demmelmair H, et al. Materno-fetal transfer of docosahexaenoic acid is impaired by gestational diabetes mellitus. American Journal of Physiology-Endocrinology and Metabolism 2013;305:E826–33. 10.1152/ajpendo.00291.2013.

[46] Daskalakis G, Marinopoulos S, Krielesi V, Papapanagiotou A, Papantoniou N, Mesogitis S, et al. Placental pathology in women with gestational diabetes. Acta Obstetricia et Gynecologica Scandinavica 2008;87:403–7. 10.1080/00016340801908783.

[47] Dasgupta S, Banerjee U, Mukhopadhyay P, Maity P, Saha S, Das B. Clinicopathological study and immunohistochemical analysis of expression of annexin A5 and apelin in human placentae of gestational diabetes mellitus. Diabetes Metab Syndr 2022;16:102435. 10.1016/j.dsx.2022.102435.

[48] Aldahmash WM, Alwasel SH, Aljerian K. Gestational diabetes mellitus induces placental vasculopathies. Environmental Science and Pollution Research 2022;29:19860–8.

[49] Karunakaran I, Nalinakumari S, Ponniraivan K. Altered Placental Histologic Features In Lean And Obese Gestational Diabetic Pregnancies. Research Journal of Pharmaceutical, Biological and Chemical Sciences 2018;9:1459–67.

[50] Whittington JR, Cummings KF, Ounpraseuth ST, Aughenbaugh AL, Quick CM, Dajani NK. Placental changes in diabetic pregnancies and the contribution of hypertension. The Journal of Maternal-Fetal & Neonatal Medicine 2022;35:486–94.

[51] Ehlers E, Talton OO, Schust DJ, Schulz LC. Placental structural abnormalities in gestational diabetes and when they develop: A scoping review. Placenta 2021;116:58–66. 10.1016/j.placenta.2021.04.005.

[52] Redline RW, Ravishankar S. Fetal vascular malperfusion, an update. APMIS 2018;126:561–9. 10.1111/apm.12849.

[53] Jaiman S, Romero R, Pacora P, Jung E, Bhatti G, Yeo L, et al. Disorders of placental villous maturation in fetal death. J Perinat Med 2020:/j/jpme.ahead-of-print/jpm-2020-0030/jpm-2020-0030.xml. 10.1515/jpm-2020-0030.

[54] Ernst LM. Maternal vascular malperfusion of the placental bed. APMIS 2018;126:551–60. 10.1111/apm.12833.

[55] Barbieri RL. One-step or two-step test for diagnosing gestational diabetes? 2021.

[56] Rudge MV, Lima CP, Damasceno DC, Sinzato YK, Napoli G, Rudge CV, et al. Histopathological placental lesions in mild gestational hyperglycemic and diabetic women. Diabetol Metab Syndr 2011;3:19. 10.1186/1758-5996-3-19.

[57] Nataly F, Hadas GH, Ohad G, Letizia S, Michal K. Is there a difference in placental pathology in pregnancies complicated with gestational diabetes A2 versus gestational diabetes A1, versus one abnormal value, on 100 gr glucose tolerance test? Placenta 2022;120:60–4. 10.1016/j.placenta.2022.02.009.

[58] The Biostatistics, Epidemiology, and Research Design Core (BERDC), University of North Dakota. School of Medicine and Health Sciences: DACCOTA Statistical Resources. University of North Dakota: School of Medicine and Health Sciences n.d. https://med.und.edu/research/daccota/berdc-resources.html (accessed March 19, 2024).

[59] Centers for Disease Control and Prevention, National Center for Health Statistics. ICD-10 - CM International Classification of Diseases, Tenth Revision, Clinical Modification (ICD-10-CM) 2023. https://www.cdc.gov/nchs/icd/icd-10-cm.htm (accessed March 19, 2024).

[60] Khong TY, Mooney EE, Ariel I, Balmus NCM, Boyd TK, Brundler M-A, et al. Sampling and Definitions of Placental Lesions Amsterdam Placental Workshop Group Consensus Statement. Archives of Pathology & Laboratory Medicine 2016;140:698–713. 10.5858/arpa.2015-0225-CC.

[61] Faye-Petersen OM, Ernst LM. Maternal Floor Infarction and Massive Perivillous Fibrin Deposition. Surgical Pathology Clinics 2013;6:101–14. 10.1016/j.path.2012.10.002.

[62] Islam MR. Sample size and its role in Central Limit Theorem (CLT). Computational and Applied Mathematics Journal 2018;4:1–7.

[63] Dinno A. Nonparametric pairwise multiple comparisons in independent groups using Dunn’s test. Stata Journal 2015;15:292–300. 10.1177/1536867x1501500117.

[64] Bewick V, Cheek L, Ball J. Statistics review 9: One-way analysis of variance. Critical Care 2004;8:130. 10.1186/cc2836.

[65] Bewick V, Cheek L, Ball J. Statistics review 10: further nonparametric methods. Critical Care 2004;8:196–9. 10.1186/cc2857.

[66] Franke TM, Ho T, Christie CA. The chi-square test: often used and more often misinterpreted. American Journal of Evaluation 2012;33:448–58. 10.1177/1098214011426594.

[67] Ford C. Pairwise comparisons of proportion. University of Virginia Library 2016. https://library.virginia.edu/data/articles/pairwise-comparisons-of-proportions (accessed April 18, 2025).

[68] Higdon R. Multiple hypothesis testing. Encyclopedia of Systems Biology, Springer, New York, NY; 2013, p. 1468–9. 10.1007/978-1-4419-9863-7_1211.

[69] Williamson T, Eliasziw M, Fick GH. Log-binomial models: exploring failed convergence. Emerg Themes Epidemiol 2013;10:14. 10.1186/1742-7622-10-14.

[70] UCLA Statistical Methods and Data Analytics. Poisson regression | r data analysis examples. UCLA Statistical Methods and Data Analytics 2021. https://stats.oarc.ucla.edu/r/dae/poisson-regression/ (accessed May 15, 2024).

[71] Lumley T, Scott A. Partial likelihood ratio tests for the Cox model under complex sampling. Statistics in Medicine 2013;32:110–23. 10.1002/sim.5492.

[72] Lumley T, Scott A. Tests for Regression Models Fitted to Survey Data. Aust N Z J Stat 2014;56:1–14. 10.1111/anzs.12065.

[73] R Documentation. Wald test for a term in a regression model. R: Analysis of Complex Survey Samples n.d. https://search.r-project.org/CRAN/refmans/survey/html/regTermTest.html.

[74] R Core Team. R: A Language and Environment for Statistical Computing 2024.

[75] Hiden U, Lassance L, Tabrizi NG, Miedl H, Tam-Amersdorfer C, Cetin I, et al. Fetal Insulin and IGF-II Contribute to Gestational Diabetes Mellitus (GDM)-Associated Up-Regulation of Membrane-Type Matrix Metalloproteinase 1 (MT1-MMP) in the Human Feto-Placental Endothelium. The Journal of Clinical Endocrinology & Metabolism 2012;97:3613–21. 10.1210/jc.2012-1212.

[76] Kadivar M, Khamseh ME, Malek M, Khajavi A, Noohi AH, Najafi L. Histomorphological changes of the placenta and umbilical cord in pregnancies complicated by gestational diabetes mellitus. Placenta 2020;97:71–8. 10.1016/j.placenta.2020.06.018.

[77] Roseboom TJ, Painter RC, de Rooij SR, van Abeelen AFM, Veenendaal MVE, Osmond C, et al. Effects of famine on placental size and efficiency. Placenta 2011;32:395–9. 10.1016/j.placenta.2011.03.001.

[78] Alwasel SH, Abotalib Z, Aljarallah JS, Osmond C, Alkharaz SM, Alhazza IM, et al. Changes in Placental Size during Ramadan. Placenta 2010;31:607–10. 10.1016/j.placenta.2010.04.010.

[79] Eriksson JG, Kajantie E, Osmond C, Thornburg K, Barker DJ. Boys live dangerously in the womb. American Journal of Human Biology 2010;22:330–5. 10.1002/ajhb.20995.

[80] Murphy VE, Gibson PG, Giles WB, Zakar T, Smith R, Bisits AM, et al. Maternal asthma is associated with reduced female fetal growth. Am J Respir Crit Care Med 2003;168:1317–23. 10.1164/rccm.200303-374OC.

[81] Paranavitana L, Walker M, Chandran AR, Milligan N, Shinar S, Whitehead CL, et al. Sex differences in uterine artery Doppler during gestation in pregnancies complicated by placental dysfunction. Biol Sex Differ 2021;12:19. 10.1186/s13293-021-00362-7.

[82] Meakin AS, Cuffe JSM, Darby JRT, Morrison JL, Clifton VL. Let’s Talk about Placental Sex, Baby: Understanding Mechanisms That Drive Female- and Male-Specific Fetal Growth and Developmental Outcomes. Int J Mol Sci 2021;22:6386. 10.3390/ijms22126386.

[83] Clifton VL. Review: Sex and the Human Placenta: Mediating Differential Strategies of Fetal Growth and Survival. Placenta 2010;31:S33–9. 10.1016/j.placenta.2009.11.010.

[84] Salafia CM, Misra DP, Yampolsky M, Charles AK, Miller RK. Allometric metabolic scaling and fetal and placental weight. Placenta 2009;30:355–60. 10.1016/j.placenta.2009.01.006.

[85] Christians JK, Chow NA. Are there sex differences in fetal growth strategies and in the long-term effects of pregnancy complications on cognitive functioning? Journal of Developmental Origins of Health and Disease 2022;13:766–78. 10.1017/S2040174422000204.

[86] Nelson SM, Coan PM, Burton GJ, Lindsay RS. Placental Structure in Type 1 Diabetes: Relations to fetal insulin, leptin, and and IGF-1. Diabetes 2009;58:2634–41. 10.2337/db09-0739.

[87] Jauniaux E, Burton GJ. Villous Histomorphometry and Placental Bed Biopsy Investigation in Type I Diabetic Pregnancies. Placenta 2006;27:468–74. 10.1016/j.placenta.2005.04.010.

[88] Higgins M, Felle P, Mooney EE, Bannigan J, McAuliffe FM. Stereology of the placenta in type 1 and type 2 diabetes. Placenta 2011;32:564–9. 10.1016/j.placenta.2011.04.015.

[89] Redline RW, Sanjita R, Bagby CM, Saab ST, Shabnam Z. Four major patterns of placental injury: a stepwise guide for understanding and implementing the 2016 Amsterdam consensus. Modern Pathology 2021;34:1074–92. 10.1038/s41379-021-00747-4.

[90] Siassakos D, Bourne I, Sebire N, Kindinger L, Whitten SM, Battaglino C. Abnormal placental villous maturity and dysregulated glucose metabolism: implications for stillbirth prevention. Journal of Perinatal Medicine 2022;50:763–8. 10.1515/jpm-2021-0579.

[91] Loukeris K, Sela R, Baergen RN. Syncytial Knots as a Reflection of Placental Maturity: Reference Values for 20 to 40 Weeks’ Gestational Age. Pediatr Dev Pathol 2010;13:305–9. 10.2350/09-08-0692-OA.1.

[92] Alqudah A. FKBPL and SIRT-1 Are Downregulated by Diabetes in Pregnancy Impacting on Angiogenesis and Endothelial Function. Frontiers in Endocrinology 2021;12.

[93] Arshad R, Kanpurwala MA, Karim N, Hassan JA. Effects of Diet and Metformin on placental morphology in Gestational Diabetes Mellitus. Pak J Med Sci 2016;32:1522–7. 10.12669/pjms.326.10872.

[94] El Sawy NAE, Iqbal MS, Alkushi AG. Histomorphological Study of Placenta in Gestational Diabetes Mellitus. Int J Morhopl 2018;36:687–92.

[95] Kamal MA, AL-Habib MFM, Selman MO. Gestational diabetes induced changes in the expression of IGF-1 in placental tissue histomorphometrical and immunohistochemical. Biochem Cell Arch 2020;20:3793–801.

[96] Parks WT. Increased Syncytial Knot Formation. In: Khong TY, Mooney EE, Nikkels PGJ, Morgan TK, Gordijn SJ, editors. Pathology of the Placenta: A Practical Guide, Cham: Springer International Publishing; 2019, p. 131–7. 10.1007/978-3-319-97214-5_17.

[97] Bhattacharjee D, Mondal SK, Garain P, Mandal P, Ray RN, Dey G. Histopathological study with immunohistochemical expression of vascular endothelial growth factor in placentas of hyperglycemic and diabetic women. J Lab Physicians 2017;9:227–33. 10.4103/JLP.JLP_148_16.

[98] Higgins M, McAuliffe F, Mooney E. Delayed villous maturation of the placenta-association with diabetes and perinatal death. American Journal of Obstetrics & Gynecology 2007;197:S111. 10.1016/j.ajog.2007.10.378.

[99] Treacy A, Higgins M, Kearney JM, Mcauliffe F, Mooney EE. Delayed Villous Maturation of the Placenta: Quantitative Assessment in Different Cohorts. Pediatr Dev Pathol 2013;16:63–6. 10.2350/12-06-1218-OA.1.

[100] Higgins M, McAuliffe FM, Mooney EE. Clinical Associations with a Placental Diagnosis of Delayed Villous Maturation: A Retrospective Study. Pediatr Dev Pathol 2011;14:273–9. 10.2350/10-07-0872-OA.1.

[101] Liang X, Zhang J, Wang Y, Wu Y, Liu H, Feng W, et al. Comparative study of microvascular structural changes in the gestational diabetic placenta. Diabetes and Vascular Disease Research 2023;20:14791641231173627. 10.1177/14791641231173627.

[102] Redline RW. Extending the Spectrum of Massive Perivillous Fibrin Deposition (Maternal Floor Infarction). Pediatr Dev Pathol 2021;24:10–1. 10.1177/1093526620964353.

